# Efficacy of face-masks used in Uganda: a laboratory-based inquiry during the COVID-19 pandemic

**DOI:** 10.1101/2020.09.28.20202952

**Authors:** Gerald Mboowa, Derrick Semugenze, Hellen Nakabuye, Douglas Bulafu, Dickson Aruhomukama

## Abstract

**Background:** With shortages of face-masks continuing to be reported worldwide, critical questions like whether or not there is an adequate alternative to commercially manufactured face-masks continue to linger especially in low- and middle-income settings. This study aimed at addressing this through testing and comparing various materials and forms of face-masks for filtration efficiency, breathability, microbial cleanliness, distance-dependent fitness, and re-usability of different face-masks procured from face-mask vendors in Kampala, Uganda.

**Methods:** This was a laboratory-based descriptive study that applied new protocols and already existing protocols with substantive modifications to ten different types of face-mask types each in quadruplicate to achieve each specified aim.

**Results:** Surgical face-masks had better filtration efficiency, distance-dependent fitness and breathability compared to other face-masks tested. Decontamination of these face-masks with 70% ethanol negatively affected their efficacy. Locally-made double layered face-masks had better: filtration efficiency, distance-dependent fitness and breathability compared to other locally-made cloth face-masks, and re-usability compared to all the face-mask types that had been tested.

**Discussion/conclusions:** Locally-made double layered cloth face-masks could serve as alternative face-masks especially for populations in low- and middle-income settings like Uganda while allowing restricted use of surgical face-masks and other respirators like the KN95 to high-risk groups only.

## Introduction

Several countries continue to encourage and/or enforce the wearing of face-masks in public settings as a preventive measure to curb the acquisition and spread of the coronavirus disease 2019 (COVID-19) that is caused by the Severe Acute Respiratory Syndrome Coronavirus 2 (SARS-CoV-2) (1–5). The use of face-masks in public settings has been reported as a crucial preventive measure in limiting the acquisition and spread of COVID-19 by the World Health Organization (WHO) and the United States Centers for Disease Control (CDC) (6,7).

Several published studies have shown that the use of face-masks in public settings can limit the acquisition and spread of COVID-19, and has several advantages for example: (i) strong sustainability, (ii) good health- and economic-benefits, and (iii) simple operation (8–15). Other published studies have described face-masks as discernable indications of the widely prevalent SARS-CoV-2, and as apparatuses that might be exploited to remind the public of the importance of the other infection-control measures for example maintaining social distancing and performing hand hygiene, and have also recommended their use to limit the acquisition and spread of COVID-19 (16). In the same study (16), face-masks have been described as symbols, beyond them being apparatuses, face-masks have been described as talismans that can boost health-care workers’ perceived sense of safety, well-being, and trust in their health-care settings.

Although testing and contact tracing remain crucial in limiting the acquisition and spread of COVID-19, designing, manufacturing and getting the public to wear face-masks seems more achievable (17–20). Hence, the use of face-masks in public settings could be the single most important-low hanging opportunity for limiting the acquisition and spread of COVID-19 and could give the public the guarantee they yearn for to return our societies back to life (17–20). Face-masks meant for use in public settings should have good qualities for example a high filtration efficiency similar to those of the highly effective respirators i.e. the N95 and KN95 respirators; face-masks equivalent to these respirators may perhaps allow the public control over their own safety, a greater incentive to use them, as well as the confidence to resume their day-to-day activities (14,17).

The design of the face-masks for use in public settings should allow for: (i) protection that could result from filtration and/or deflection i.e. how well potentially infectious aerosols are blocked from passing through the face-masks as well as “fit” and leakage i.e. how well the face-masks seal around the wearer’s face and prevent potentially infectious aerosols from coming around them, while permitting breathed out air to vent efficiently (17,21,22), (ii) scalability that is permitted by the design’s potential to utilize commonly available materials that both commercial and non-commercial manufacturers can procure in huge quantities (17), (iii) comfort that allows the wearers to use the face-masks for extended durations of time without having to touch them or take them off too frequently (17,23), (iv) re-usability to prevent the continuous need for new face-masks; face-masks should be easy to clean, this would enable their repeated use (17,19,23), (v) style since the wide-spread adoption of face-mask use could necessitate a significant cultural shift so that they could become a seamless part of a “new normal”, could be enhanced by the use of various logos and/or colors of individuals’ favorite sports teams and/or brands (17,23). However, it is critical to note that designing and manufacturing such face-masks and influencing the public to use them as one of the preventive measures to curb the acquisition and spread of COVID-19 is not straight forward as it poses not only engineering and manufacturing challenges but also marketing challenges that could necessitate quite a lot of tradeoffs (17).

Also critical to note is that, not only did the outbreak of COVID-19 spark a major debate about the efficacy of face-masks in general but it also exposed the inadequacy of existing guidelines on the use of cloth face-masks, despite the wide spread use of these face-masks in majority of the low- and middle-income countries especially in Africa as reported in several studies (20,24–27). At the same time, the outbreak fashioned critical questions like whether or not there was an adequate alternative to commercially manufactured face-masks for example the N95, KN95 and surgical face-masks that could be made easily available for populations in low- and middle-income settings especially in Africa (28). At the time of writing, similar to a few other low- and middle-income countries in Africa, Uganda was continuing to implement a phased-approach of lifting the countrywide lockdown while considering the use of face-masks in public settings mandatory for all (29). In light of this, many untested brands of cloth face-masks, potentially inadequate to provide protection against COVID-19 could have flooded the Ugandan market hence making it necessary to assess the safety and fitness-for-use of the different brands of cloth face-masks that were commonly circulating on the Ugandan market during the COVID-19 pandemic.

Hence this study aimed at testing and comparing various materials and forms of cloth face-masks procured from face-mask vendors in Kampala, Uganda during the COVID-19 pandemic; the testing was done to assess the: filtration efficiency, breathability, microbial cleanliness, distance-dependent fitness, and re-usability of the cloth face-masks. We hoped that the findings obtained from this study would assist health organizations in Uganda and similar low- and middle-income settings in Africa to develop policy on the use of cloth face-masks to prevent the transmission of COVID-19.

## Results

### Filtration efficiency of the face-masks tested

All the face-masks that had been tested had a filtration efficiency of ≥ 99.9%, regardless of the face-mask decontamination method used (Table 1).

**Table 1.**
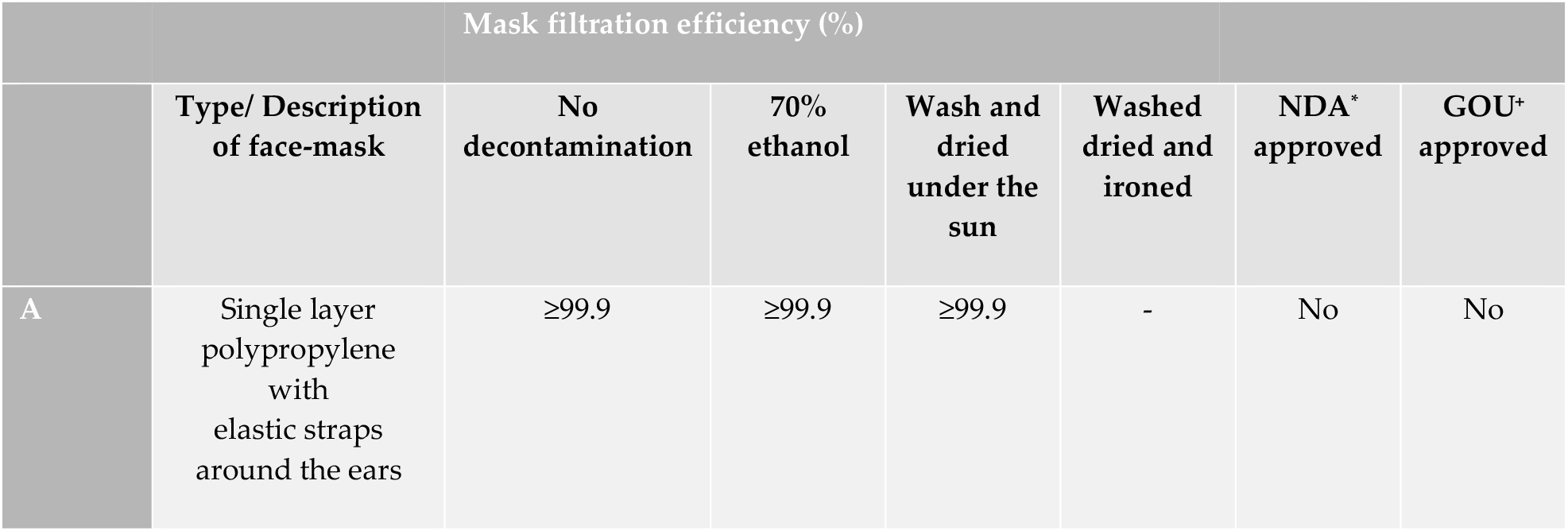
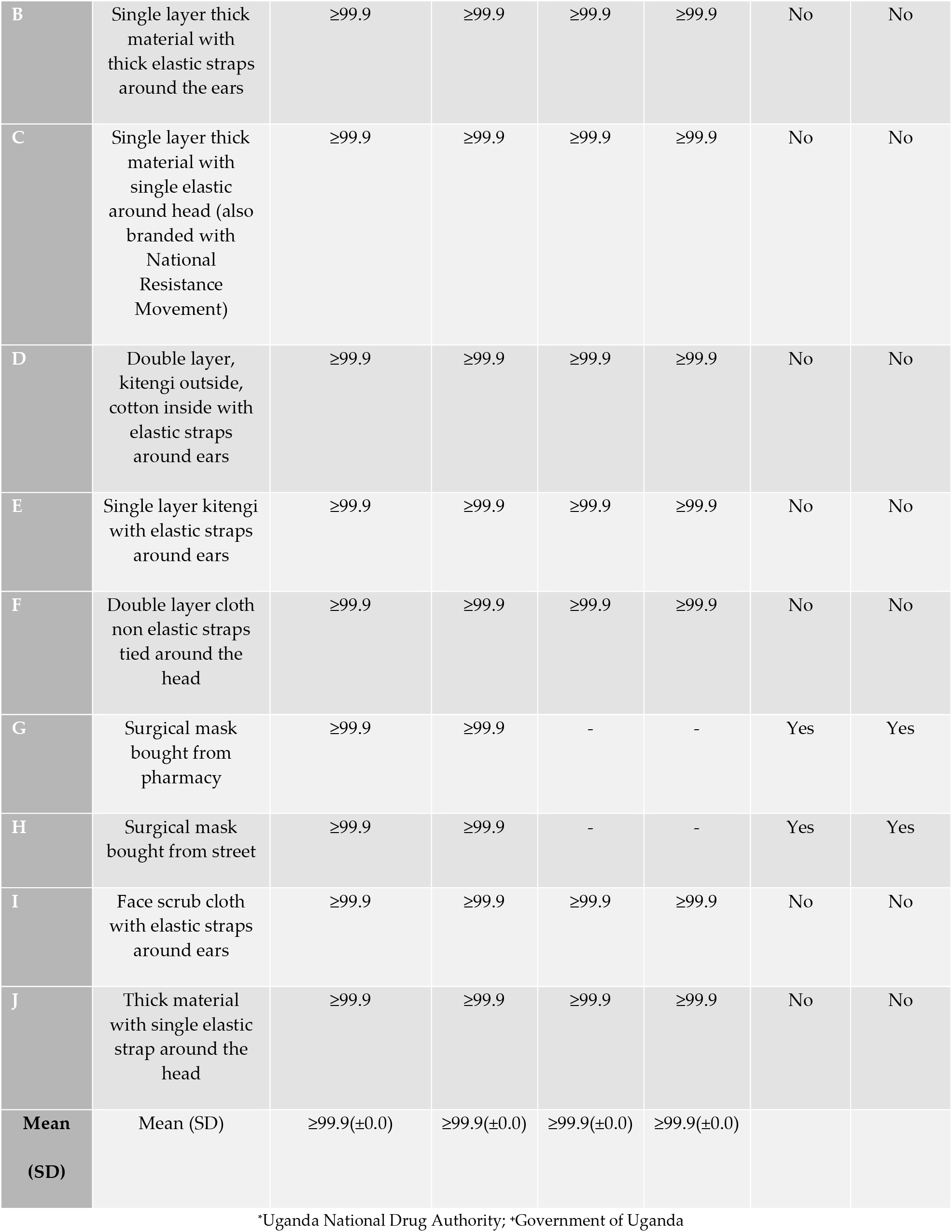
Filtration efficiency of the face-masks.

| Mask filtration efficiency (%) |  |  |  |  |  |  |  |
| --- | --- | --- | --- | --- | --- | --- | --- |
|  | Type/ Description of face-mask | No decontamination | 70% ethanol | Wash and dried under the sun | Washed dried and ironed | NDA* approved | GOU* approved |
| A | Single layer polypropylene with elastic straps around the ears | $\geq 99.9$ | $\geq 99.9$ | $\geq 99.9$ | - | No | No |
| B | Single layer thick material with thick elastic straps around the ears | ≥99.9 | ≥99.9 | ≥99.9 | ≥99.9 | No | No |
| C | Single layer thick material with single elastic around head (also branded with National Resistance Movement) | ≥99.9 | ≥99.9 | ≥99.9 | ≥99.9 | No | No |
| D | Double layer, kitengi outside, cotton inside with elastic straps around ears | ≥99.9 | ≥99.9 | ≥99.9 | ≥99.9 | No | No |
| E | Single layer kitengi with elastic straps around ears | ≥99.9 | ≥99.9 | ≥99.9 | ≥99.9 | No | No |
| F | Double layer cloth non elastic straps tied around the head | ≥99.9 | ≥99.9 | ≥99.9 | ≥99.9 | No | No |
| G | Surgical mask bought from pharmacy | ≥99.9 | ≥99.9 | - | - | Yes | Yes |
| H | Surgical mask bought from street | ≥99.9 | ≥99.9 | - | - | Yes | Yes |
| I | Face scrub cloth with elastic straps around ears | ≥99.9 | ≥99.9 | ≥99.9 | ≥99.9 | No | No |
| J | Thick material with single elastic strap around the head | ≥99.9 | ≥99.9 | ≥99.9 | ≥99.9 | No | No |
| **Mean (SD)** | Mean (SD) | ≥99.9(±0.0) | ≥99.9(±0.0) | ≥99.9(±0.0) | ≥99.9(±0.0) |  |  |

\*Uganda National Drug Authority; \*Government of Uganda

### In-house breathability testing of the face-masks

Glass beakers that had been sealed with surgical face-masks that had been obtained from both the community pharmacy and street vendors, as well as the locally-made double-layered cloth face-masks with non-elastic straps purposely for tying around the head showed a significant increase in weight after the 24-hour duration of the testing. However, glass beakers that had been sealed with surgical face-masks that had been obtained from the street vendors had a significantly higher weight gain compared to those that had been obtained from the community pharmacy and the locally-made double-layered cloth face-masks with non-elastic straps purposely for tying around the head, while the locally-made double-layered cloth face-masks with non-elastic straps purposely for tying around the head had a significantly higher weight gain compared to the other face-masks that had been tested, regardless of the face-mask decontamination method used (Figure 1b). Face-mask pictorial (Figure 1a).

**Figure 1a.**
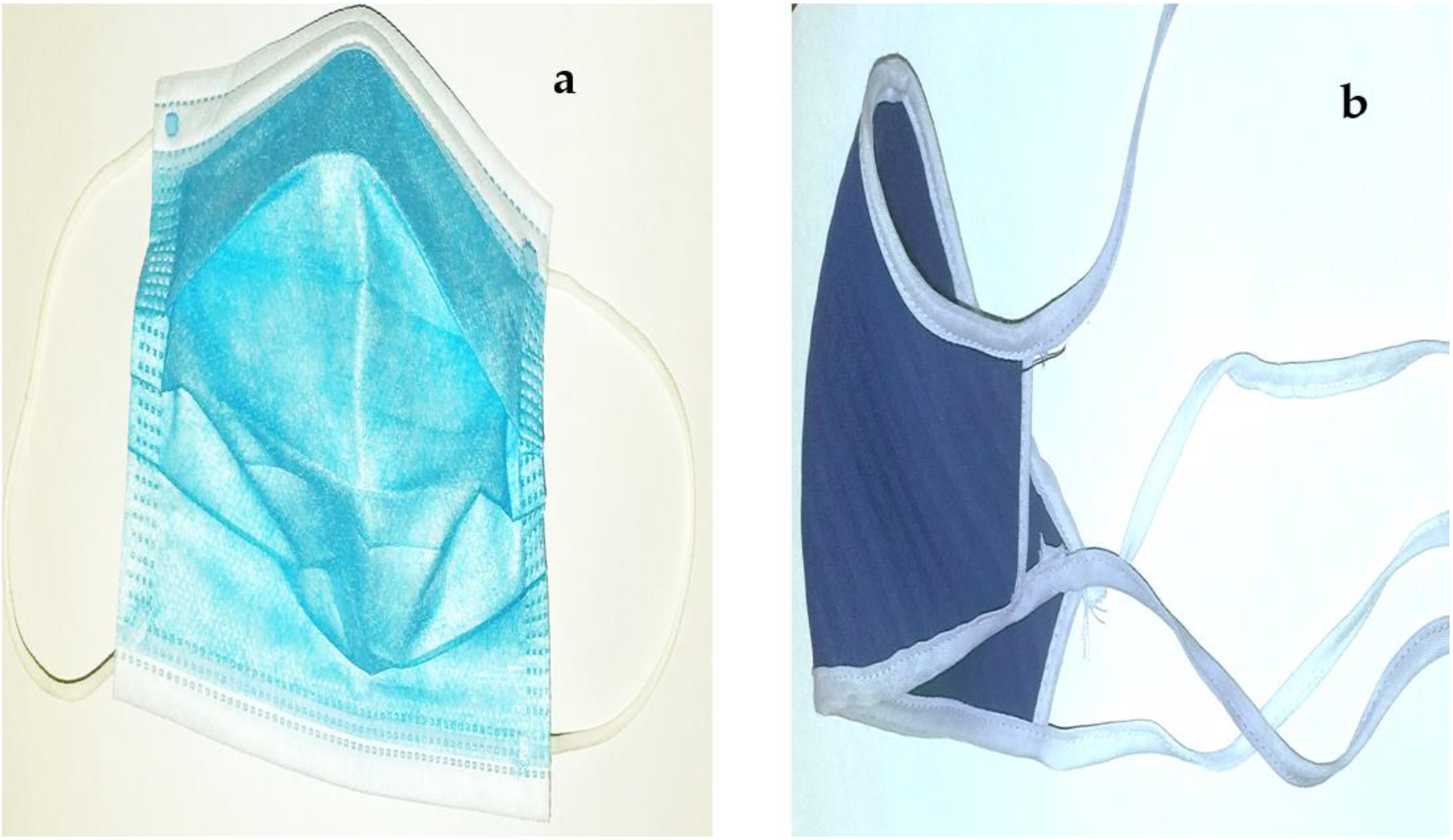
Images of: (a) a surgical face-mask obtained from the community pharmacy and (b) a locally-made double-layered cloth face-mask with non-elastic straps.

**Figure 1b.**
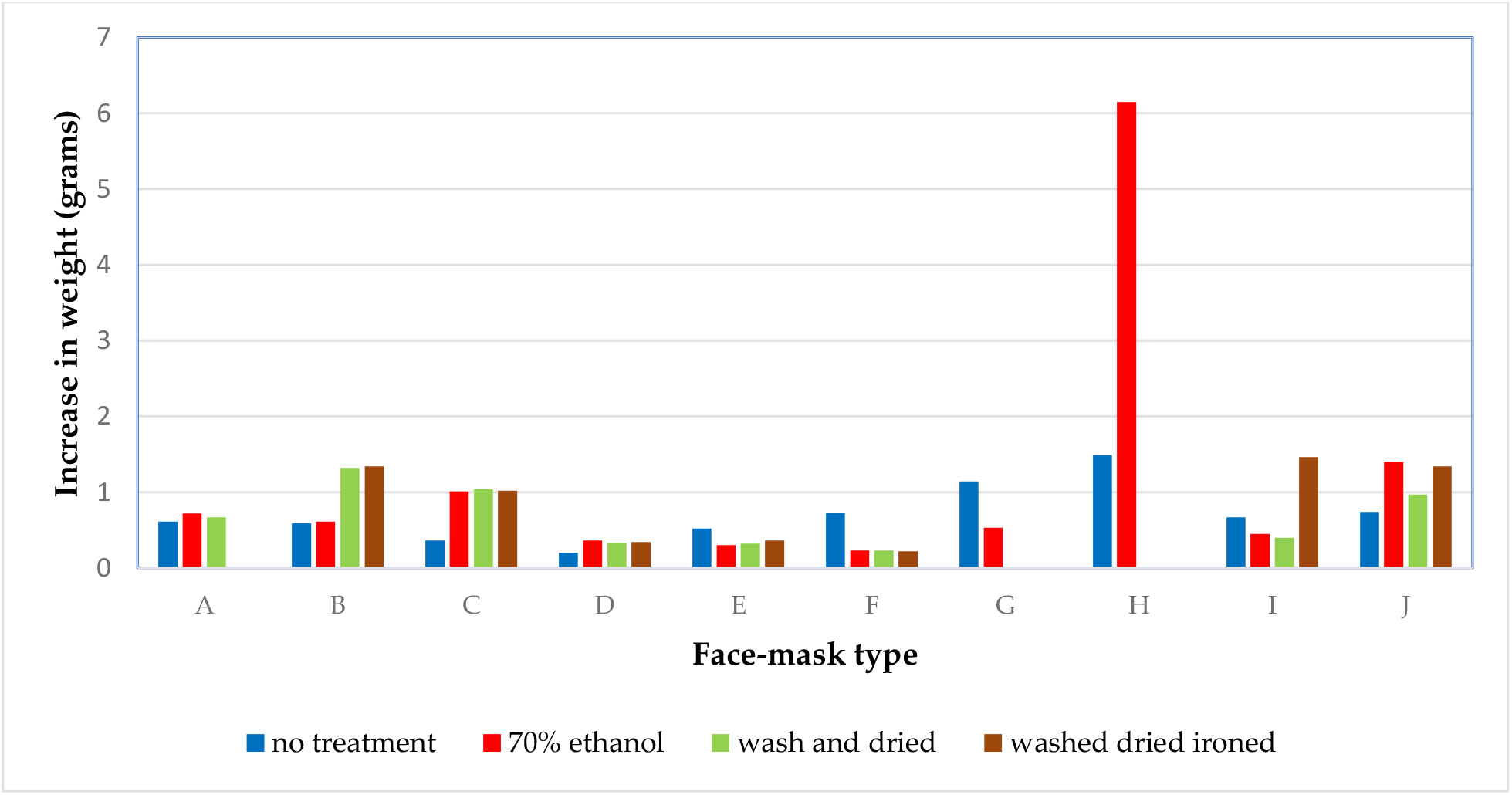
Testing for breathability.

Furthermore, there was a significant association between the type of face-mask and the average increase in weight of each of the face-masks used in the testing, regardless of the method of face-mask decontamination used (*F=4*.*42, p<0*.*05*). However, there was no significant association between the type of face-mask and the humidity content in the testing environment, regardless of the method of face-mask decontamination used (*F=1*.*16, p>0*.*05*). Surgical face-masks that had been obtained from street vendors had a significantly higher weight gain compared to the other types of face-masks that had been tested (Table 2).

**Table 2.**
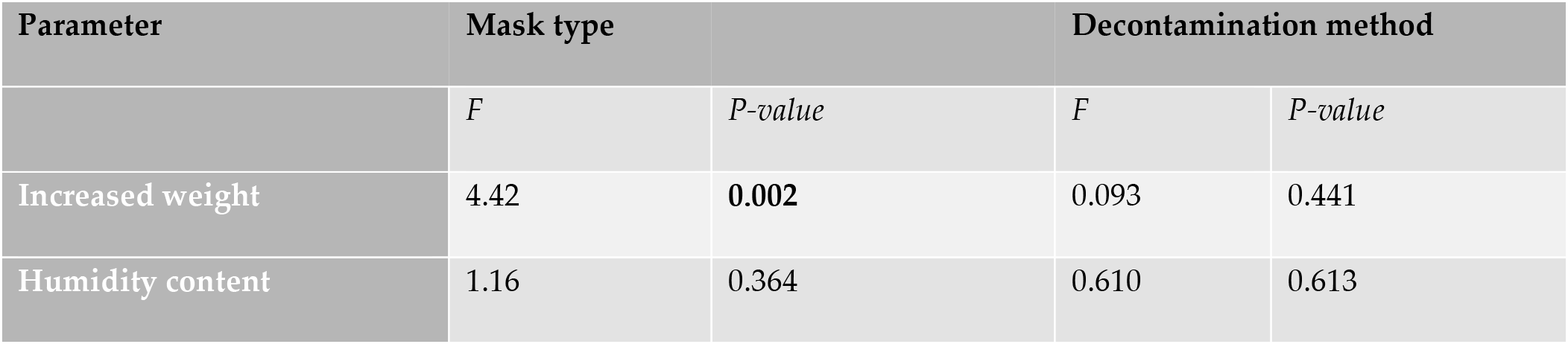
Association testing in the in-house breathability testing.

| Parameter | Mask type |  | Decontamination method |  |
| --- | --- | --- | --- | --- |
|  | *F* | *P-value* | *F* | *P-value* |
| Increased weight | 4.42 | 0.002 | 0.093 | 0.441 |
| Humidity content | 1.16 | 0.364 | 0.610 | 0.613 |

### Microbial cleanliness of the face-masks

There was no significant difference between the counts in terms of CFUs and type of microorganisms that had been isolated from the inner- and outer-surfaces of the face-masks that had been tested, regardless of the method of face-mask decontamination used (Figure 2). The gram-positive bacteria that had been isolated from both the inner- and outer-surfaces of the face-masks surfaces were predominantly coagulase negative staphylococcus (>95%), while no gram-negative bacteria had been isolated.

**Figure 2.**
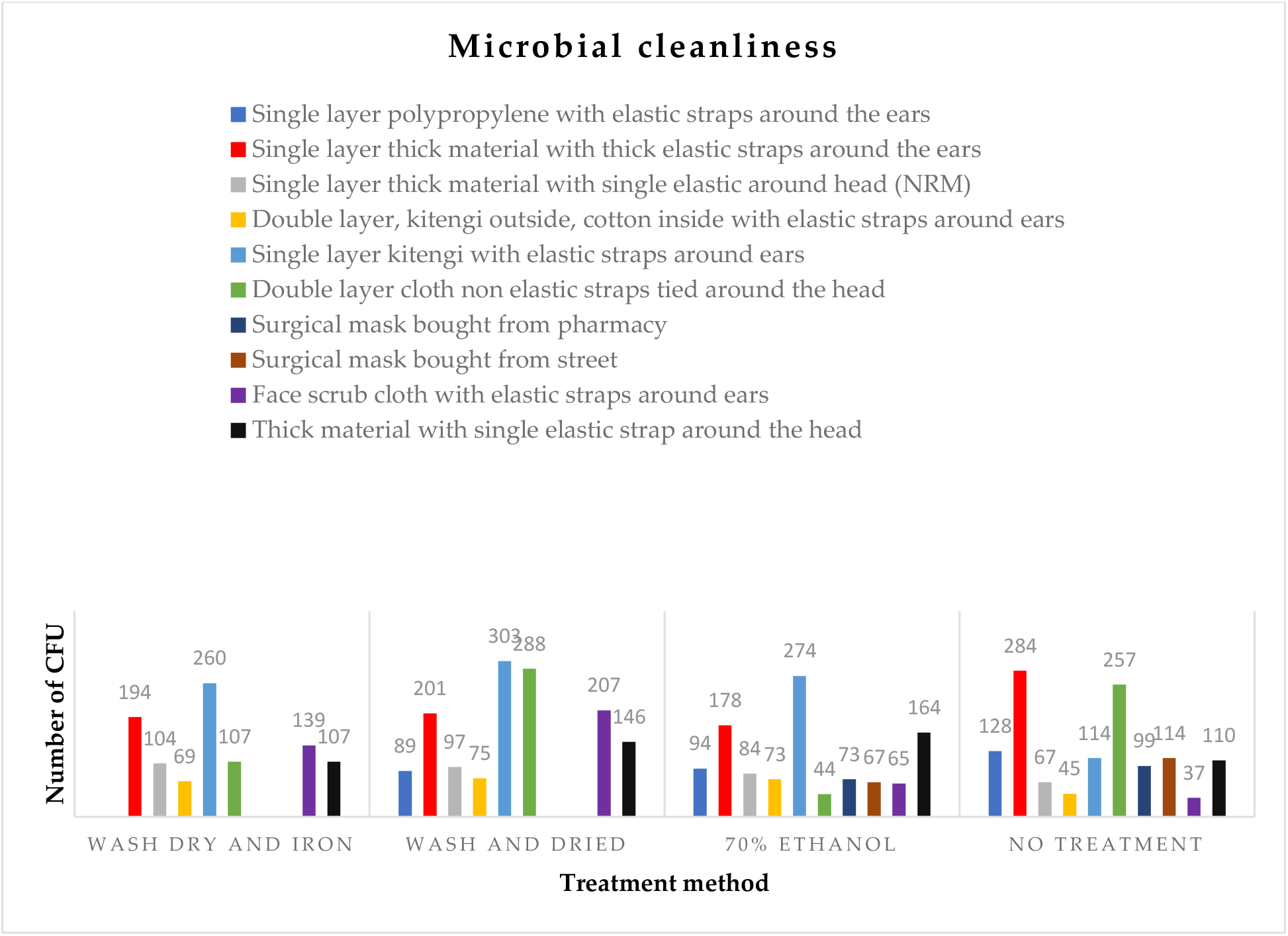
Counts of microorganisms from face-mask surfaces upon microbiological culture.

### Distance dependent fitness testing of the face-masks

Figures 3, 4, 5, and 6 show the relationship between the number of sprays that were applied to each face-mask with distance, as the measure for distance dependent fitness testing of the face-masks. The distance dependent fitness testing experiment was done for face-masks that had not been subjected to any decontamination, those that had been: treated with 70% ethanol, washed and dried under the sun and washed, dried under the sun and ironed.

### With no decontamination

Largely, the number of sprays applied to each of the face-masks for the volunteers to taste the reagent that had been used in the testing increased with increase in the distance. At 1 meter, the study volunteers tasted the reagent that had been used in the testing after just a few sprays (<100 sprays). As the distance increased, the number of sprays applied to each of the face-masks also increased. Noteworthy, the number of sprays applied in the case of the surgical face-masks that had been obtained from the street vendors for the volunteers to taste the reagent used in the testing increased exponentially to ≥ 500 sprays at 2 meters. The number of sprays applied to the locally-made double-layered cloth face-masks with non-elastic straps purposely for tying around the head for the volunteers to taste the reagent used in the testing also increased steadily till 2 meters at which point they increased exponentially passing the 500-spray mark at 3 meters. The number of sprays applied to the face-mask that had been made from thick material with an elastic strap for attaching around the head for the volunteers to taste the reagent used in the testing also increased steadily till 5 meters at which point they passed the 500-spray mark. The number of sprays applied to all the single layered face-masks for the volunteers to taste the reagent used in the testing increased steadily till 6 meters but did not pass the 500-spray mark.

The surgical face-masks that had been bought from the street vendors passed the distance dependent test at 2 meters, while the locally-made double-layered cloth face-masks passed the same test at 3 meters, and the face-masks that were made from the thick material with an elastic strap for attaching around the head passed the same test at 6 meters. All single-layered face-masks did not pass for distance dependent fitness test.

**Figure 3.**
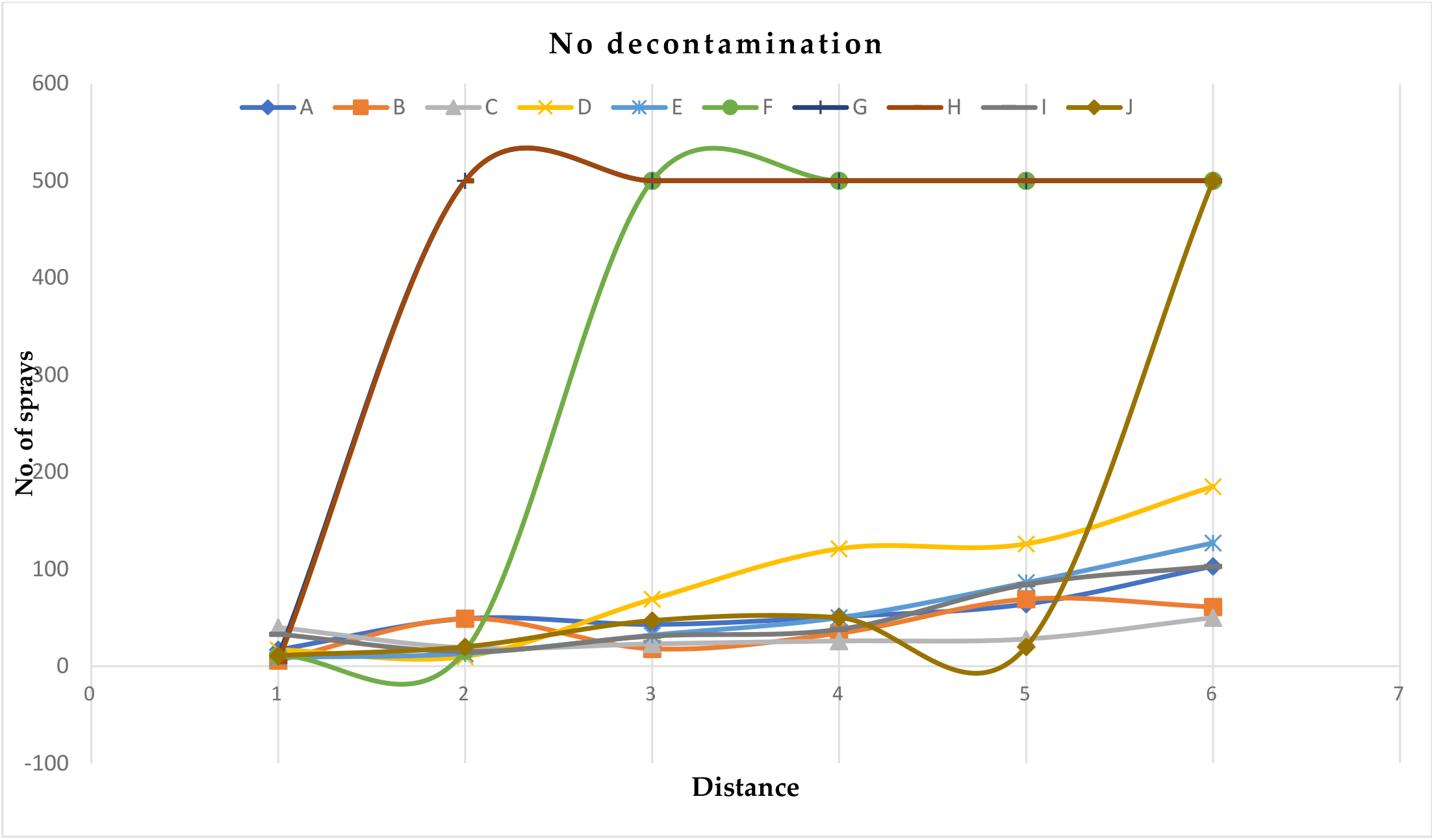
Distance dependent fitness testing of the face-masks without decontamination.

### Decontamination with 70% ethanol

Largely, the number of sprays applied to each of the face-masks for the volunteers to taste the reagent that had been used in the testing increased with increase in the distance upon decontamination with 70% ethanol. At 1 meter, all the face-masks required a few sprays (< 100 sprays) to be applied for the volunteers to taste the reagent that had been used in the testing. As the distance increased, the number of sprays applied to the locally-made double-layered cloth face-masks with non-elastic straps purposely for tying around the head for the volunteers to taste the reagent used in the testing increased steadily till 2 meters at which point they increased exponentially passing the 500-spray mark at 3 meters. The number of spays applied to face-masks that had been made of face scrub cloth with elastic straps that allowed for attachment on the ears for the volunteers to taste the reagent that had been used in the testing increased steadily till 5 meters at which point they then increased exponentially passing the 500 spray-mark at 6 meters.

On decontamination with 70% ethanol, the locally-made double-layered cloth face-masks with non-elastic straps purposely for tying around the head, and face-masks that had been made of face scrub cloth with elastic straps that allowed for attachment on the ears passed the distance dependent fitness test. Noteworthy, both surgical face-masks that had been obtained from the community pharmacy and street vendors failed the distance dependent fitness test.

**Figure 4.**
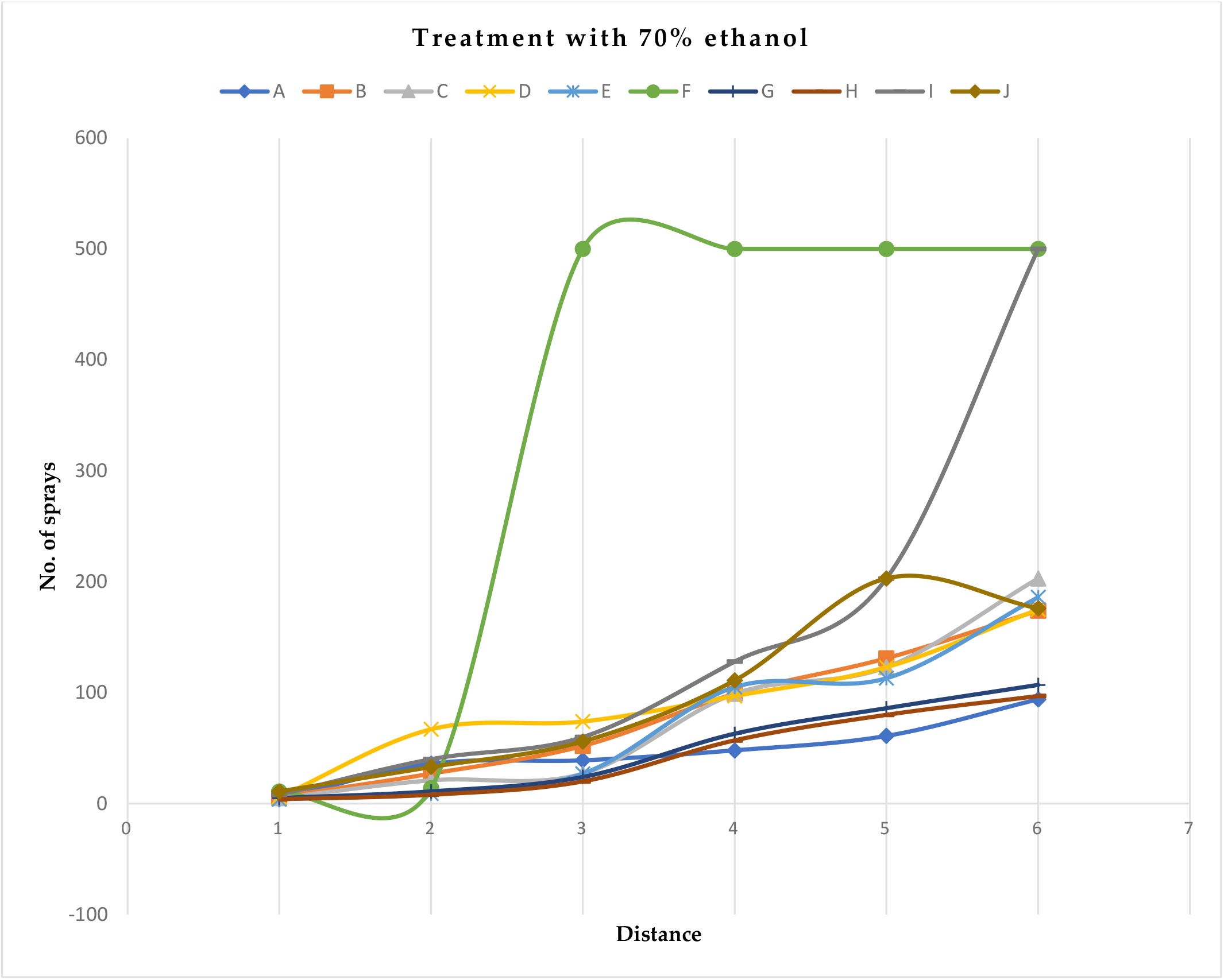
Distance dependent fitness testing of the face-masks after decontamination with 70% ethanol.

### Washing of face-masks with non-bacterial soap and drying them under the sun

Upon this face-masks decontamination, the number of sprays applied to each of the face-masks for the volunteers to taste the reagent that had been used in the testing largely increased with increase in the distance. At 1 meter, all the face-masks required a few sprays (<100 sprays) to be applied for the volunteers to taste the reagent that had been used in the testing. However, as the distance increased, the number of sprays applied to the locally-made double-layered cloth face-masks with non-elastic straps purposely for tying around the head for the volunteers to taste the reagent used in the testing increased steadily till 3 meters at which point they increased exponentially till they passed the 500 spray-mark at 4 meters. The number of sprays applied to the face-masks that had been made of face scrub cloth with elastic straps that allowed for attachment on the ears, steadily increased till 5 meters where they then increased exponentially passing the 500 spray-mark at 6 meters.

Upon washing of the face-masks with non-bacterial soap and drying them under the sun: the locally-made double-layered cloth face-masks with non-elastic straps purposely for tying around the head and face-masks made of face scrub cloth with elastic straps for attaching onto the ears passed the distance dependence fitness test.

**Figure 5.**
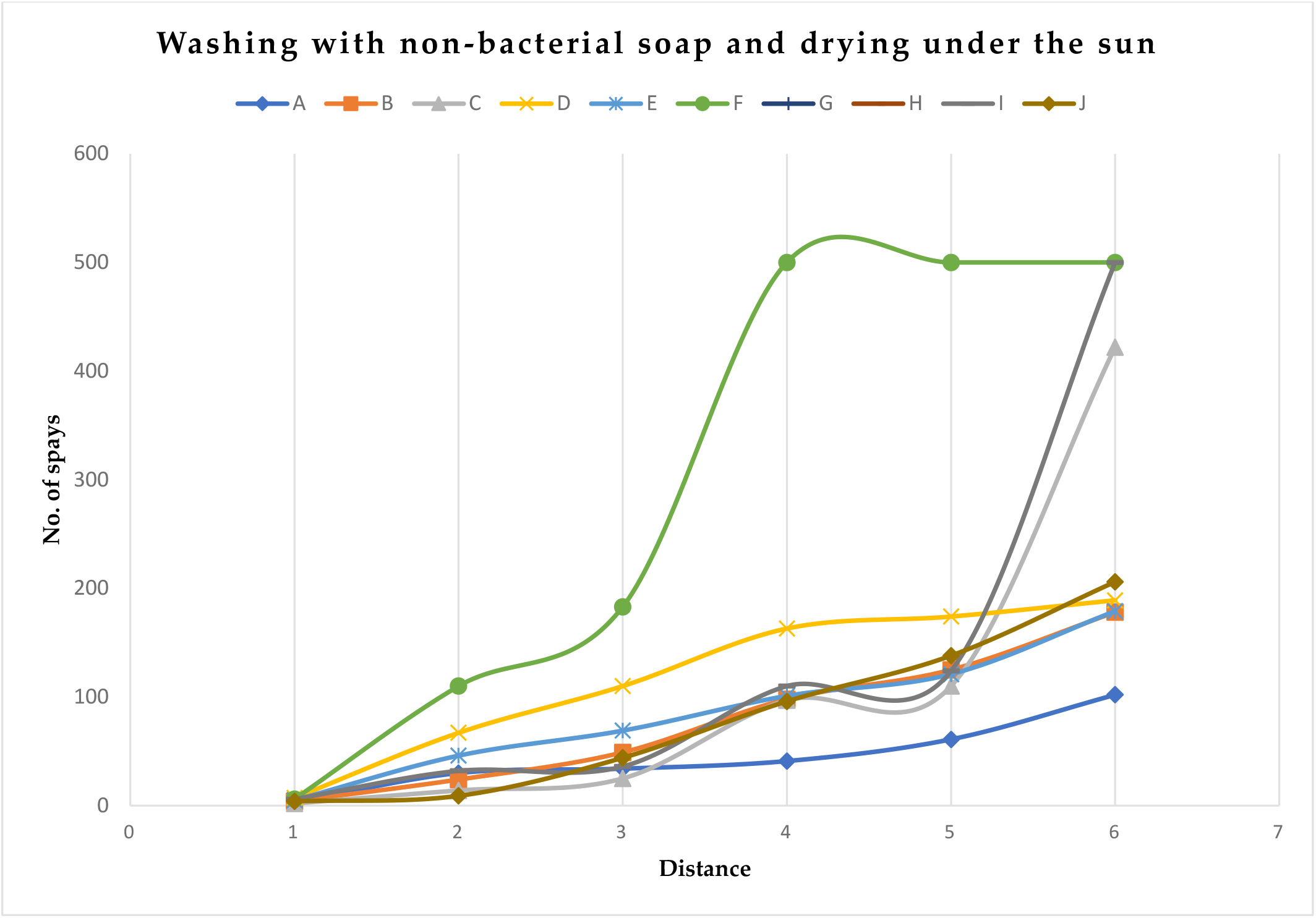
Distance dependent fitness testing of the face-masks after washing them with non-bacterial soap and drying them under the sun.

### Washing of face-masks with non-bacterial soap, drying them under the sun and ironing them

Generally, the number of sprays applied to each of the face-masks for the volunteers to taste the reagent that had been used in the testing increased with increase in the distance. At 1 meter, all the face-masks required a few sprays (<100 sprays) to be applied for the volunteers to taste the reagent that had been used in the testing. As the distance increased, the number of sprays applied to the locally-made double-layered cloth face-masks with non-elastic straps purposely for tying around the head for the volunteers to taste the reagent used in the testing increased steadily till 2 meters at which point they increased exponentially passing the 500 spray-mark at 3 meters. The number of sprays applied to the face-masks made of face scrub cloth with elastic straps for attaching onto the ears increased steadily till 5 meters at which point they increased exponentially passing the 500 spray-mark at 6 meters.

On washing of face-masks with non-bacterial soap, drying them under the sun and ironing the, locally-made double-layered cloth face-masks with non-elastic straps purposely for tying around the head and the face-masks made of face scrub cloth with elastic straps for attaching onto the ears passed the distance dependent fitness test.

**Figure 6.**
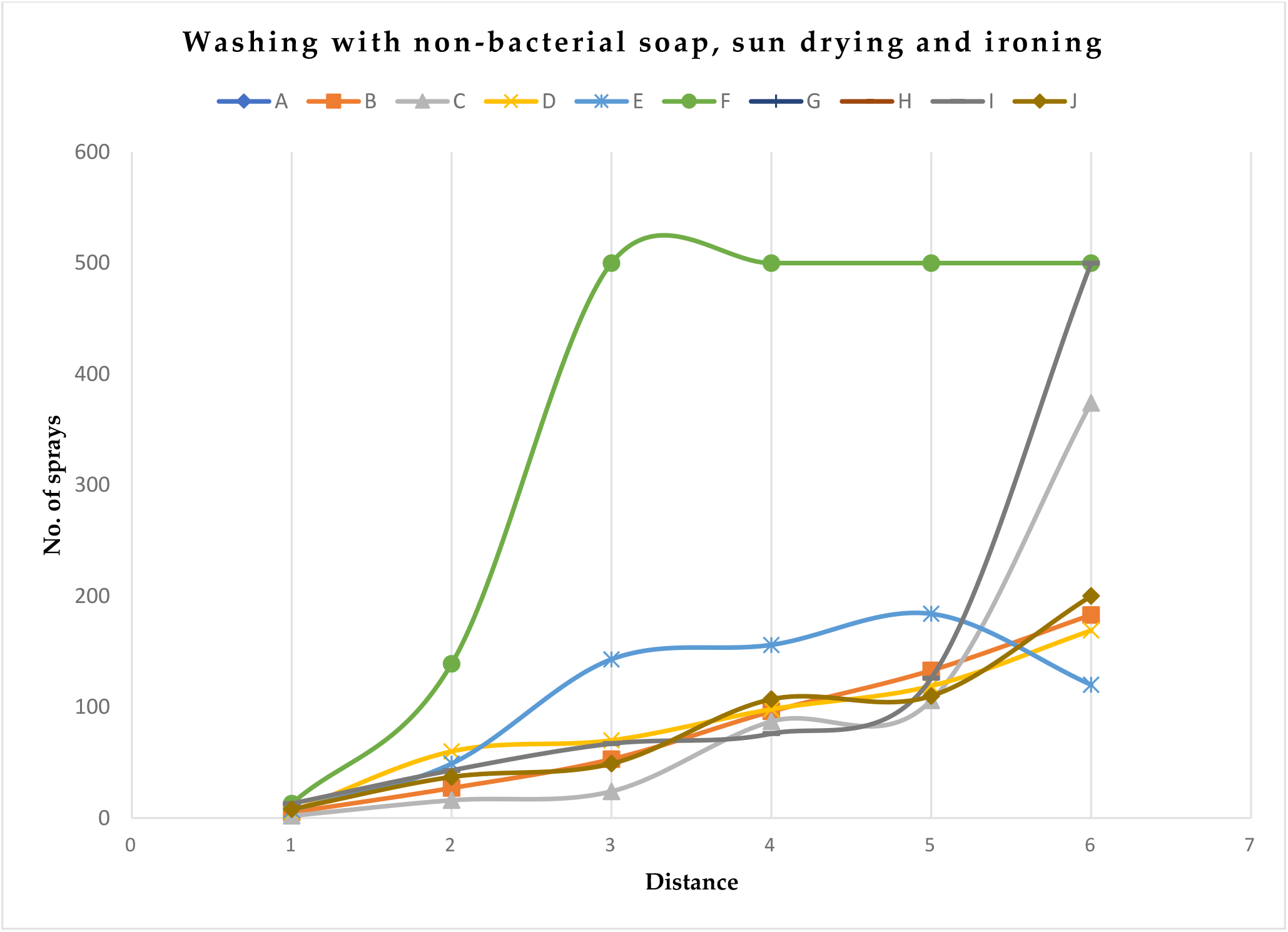
Distance dependent fitness testing of the face-masks after washing them with non-bacterial soap, drying them under the sun and ironing them.

## Discussion

To the best of our knowledge this is the first study assessing the efficacy of face-masks in Uganda. In this study, we analyzed the: filtration efficiency, breathability, microbial cleanliness, distance-dependent fitness, and re-usability of different face-masks procured from face-mask vendors in Kampala, Uganda during the COVID-19 pandemic. This study answered questions like whether or not there was an adequate alternative to imported commercially manufactured face-masks for example the KN95 and surgical face-masks, through testing and comparing various materials and forms of face-masks. The findings of this study could be beneficial for improving the uptake of face-masks by the public in Uganda and similar low- and middle-income countries in Africa, as a key intervention in preventing the transmission of COVID-19. In addition, the findings of this study unmasked the potential of a locally-made face-mask to offer an alternative to imported commercially manufactured face-masks in providing protection against COVID-19, and hence inspires efforts geared towards local production of face-masks not only in Uganda but also in other low- and middle-income countries in Africa.

The increased local-production of face-masks could translate into increased supply and availability or accessibility which could address future shortages, as well as ensure reasonable prices of the face-masks. Increased local-production could also enable prioritizing and limiting the use of KN95 and surgical face-masks to frontline clinicians and essential health-care workers.

This study’s finding where surgical face-masks purchased from either the community pharmacy or street vendors had better filtration efficiency and breathability than any other type of face-masks that had been tested are consistent with the findings of related studies. These studies (30–32), alluded to the effectiveness of surgical face-masks in offering protection against infectious aerosols for example SARS-CoV-2 while attributing this to their good qualities of filtration and breathability. However, this finding is inconsistent with those of other studies (33–35), that reported filtration limitations of surgical face-masks which could negatively affect the ability of these face-masks to offer protection to the wearers against infectious aerosols for example SARS-CoV-2 especially during coughs by infected patients.

Above and beyond the apparent imperfect filtration efficiency and breathability of surgical face-masks, several observational studies have found no significant benefits of other respirators (i.e. the N95 and KN95) over these face-masks and have indicated the ability of these face-masks to not only offer protection against infectious aerosols for example SARS-CoV-2 but to also offer a cheaper alternative to the other respirators (i.e. the N95 and KN95), which have been reported to offer better protection than the surgical face-masks (30,36–39). Therefore, despite the limitations of surgical face-masks there remains reason for optimism regarding their real-world effectiveness.

The differences in performance of the surgical face-masks that had been obtained from the community pharmacy and the street vendors reported in this study could be due to the differences in the manufacturing enterprises of these face-masks. Different manufacturing enterprises use different materials in production and this could lead to differences in the performance of the face-masks they produce. A similar line of thinking has been documented by related studies (40). In these studies, surgical face-masks were reported to provide different levels of protection against infectious aerosols particularly when they were produced from different enterprises.

The rapid growth in consumer demand for sustainable face-masks represents an enormous opportunity for small-scale enterprises in low- and middle-income countries. We believe that providing assistance to these small-scale enterprises could enable them to seize the opportunity and significantly improve the quality of the face-masks they produce. This could also consequently result in the reduction of the differences in the face-masks thereby creating more homogeneity in performance of the face-masks.

Global and local shortages of respirators and surgical face-masks have been reported by several studies (37,41). These shortages have caused the extended use or re-use of respirators as well as surgical face-masks designed for single-use (37,41–43). To ease the shortage of respirators and surgical face-masks, many individuals continue to explore non-standard methods to maintaining an adequate supply including among others face-mask decontamination, which could extend the wearable life of the face-masks beyond the expiration date, and procuring of the more costly respirators i.e. KN95 and N95 (37). However, the findings of this study indicate that the decontamination of surgical face-masks obtained from both the community pharmacy and street vendors using 70% ethanol negatively affected the general efficacy of these face-masks. These negative effects could have been due to the 70% ethanol destroying the structural integrity of the surgical face-masks, a thinking that has also been shared in related studies (43–45). These studies have also reported the potential of using other decontamination methods for example using vaporized hydrogen peroxide and ultraviolet germicidal irradiation (41,46–48). However, basing on the evidence generated in this study about the negative effects on the efficacy when decontamination of surgical face-masks is done with 70% ethanol, as well as the alternative potential decontamination approaches provided by related studies, but which have been also documented to be labor intensive and expensive (47), hence likely to be unaffordable by majority of the population and health-care systems in low-and middle-income countries, health-care organizations for example ministries of health in these countries could ensure that policies as well as systems are put in place to ensure that decontamination practices are carried out safely and in line with existing evidence. Also, these organizations could ensure that the available surgical face-masks are restricted for use to only the frontline clinicians and essential health-care workers to reduce the requirements for re-use and extended use of the surgical face-masks.

The findings in which locally made, double layered cloth face-masks were found to provide the best alternatives to surgical face-masks could be related to those of related studies (14,28). In these studies, testing of face-masks that had been made from two layers of quilt fabric with household paper towels as filters was done, the face-masks tested in these studies were reported to be viable alternatives to surgical face-masks and other respirators like the KN95 and N95 in providing protection to the users against infectious aerosols. Despite the fact that these studies acknowledged the superiority of surgical face-masks over cloth face-masks, they highlighted that the use of these face-masks was severalfold more effective than not wearing a face-mask at all.

As the world with particularly low- and middle-income settings standing to benefit the most, seek cheaper approaches of achieving mask decontamination as reported in related studies (47), this study’s findings that demonstrated the potential of decontaminating locally made, double layered cloth face-masks utilizing cheaper local methods and the ability of the locally made, double layered cloth face-masks to maintain their structural integrity and hence efficacy despite the different methods of decontamination the face-masks had been subjected to are of great importance. These findings imply, that once procured, these face-masks can be used and re-usable or used for extended durations of time while decontaminating the face-masks with the documented approaches unlike in the case of surgical face-masks. The findings could also imply that locally made, double layered cloth face-masks when availed and used could improve the uptake of face-masks by the public in Uganda and similar low- and middle-income countries in Africa, as a key intervention in preventing the transmission of COVID-19. Also, similar to other the findings of related studies (49,50), these findings reinforce the potential for local mass production of protective face-masks by likely non-experts utilizing the available resources through policy information as well as call for the updating of national guidelines to provide clear and consistent criteria to ensure the widest availability and appropriate use of face-masks, while bearing in mind populations in social disadvantaged or low- and middle-settings.

Our results demonstrated that locally made, double layered cloth face-masks can be effectively decontaminated using various approaches while maintaining their structural integrity which means maintaining their ability of provide protection to the wearer against infectious aerosols. Given the Ugandan government recommendation that cloth face-masks should be worn in public settings, decontamination using ordinary soap and drying under the sun with or without ironing could provide cheaper, safe and effective decontamination of cloth face-masks.

Regarding microbial cleanliness of the face-masks, of note we found no significant difference between the counts as well as the types of microorganisms that had been isolated from the inner- and outer surfaces of the face-masks that had been tested in this study. These finding are inconsistent with those of related studies, that reported having found a greater contamination of on the outer-surfaces of the face-masks with microorganisms as compared to the inner-surfaces of the same face-masks (35). Coagulase negative staphylococcus remain known members of the normal flora of the human skin, nasal and oral cavity (51–53). The isolation of coagulase negative staphylococcus from the inner- and outer-surfaces of face-masks in this study could be attributed to several factors that could include among others: face-masks aerodynamic features that could result in a turbulent jet due to leakage of air around the edge of the face-mask that could consequently contaminate the outer-surfaces of the face-mask, a thought shared by related studies (35), as well as the possibility that the bacteria could have: been picked or originated from the skin, oral, nasal cavities of the wearers or even their hands during contact with the face-masks (53,54). These findings hence support the importance of hand hygiene after touching the surfaces of face-masks to ensure microbiological cleanliness of the face-masks.

Our study had strength and limitations; (i) the strength of the study is that, in absence of documented protocols, the study was able to develop, test and optimize protocols to successfully perform the testing of the face-masks, protocols which could also be adapted as templates for the development of more comprehensive protocols, (ii) despite the fact that the size and concentrations of the SARS-CoV-2 in aerosols generated during coughs by infected patients are unkown, the sizes of aerosols generated by our hand-held sprayer especially at different distances were not determined, this leaves a gap in the information regarding whether or not the face-masks tested would offer adequate protection against SARS-CoV-2, (iii) most experiments done especially in the high income settings use automated aerosol generation to continuously produce aerosols in the testing, however, in this study, we used hand-held sprayers whose aerosol generation depends on the number of squeezes made by the operator, (iv) despite measuring environmental temperature and humidity since they have been documented to affect the evaporation of the generated aerosols during ordinary exhalation, talking and singing, the wind speed was not catered for during this study and is likely to have affected the aerosol transmission and speed.

## Conclusions

This study described workable protocols to evaluate the efficacy of face-masks. This study also demonstrated limitations of surgical face-masks in the context of re-usability, although they are to a higher extent protective. In addition, this study also demonstrated the potential of locally-made double layered cloth face-masks to serve as cheaper face-mask alternatives especially for populations in Uganda. Unmasking the potential of the locally-made double layered cloth face-masks allows for the limited use of surgical face-masks to high-risk groups. Lastly, the study demonstrated the need to boost local production of face-masks by small-scale enterprises with support of relevant government ministries.

## Methods

### Study design

This was a laboratory-based descriptive study, and was part of a larger study titled: Assessing knowledge, attitudes, perceptions and skills towards the use of face-masks: a community-level perspective (MASKUG-2020), that aimed to assess: (1) knowledge, attitudes, perceptions, practices, and skills towards the use of face-masks by high-risk groups in Kampala, Uganda, (2) skills towards the use of face-masks by the same groups, and (3) to evaluate the face-masks for safety and fitness-for-use, (4) to provide a classification for those commonly circulating on the Ugandan market, as well as (5) to educate and skill the same groups on the rational use and disposal of face-masks.

### Study sites

The study was carried out in the Department of Medical Microbiology at the School of Biomedical Sciences, College of Health Sciences, Makerere University-Uganda.

### Description and source of the face-masks

Ten different types of face-masks each in quadruplicate, were purposively selected and procured from face-mask vendors in Kampala, the capital city of Uganda (Table 1).

### Laboratory testing

This was achieved using new protocols as well as already existing protocols with substantive modifications or rather innovative applications of existing protocols to new models or scientific questions and involved;

### Semi-quantitative mask fitness testing

This was performed using saccharin solution. Saccharin is a non-nutritive sweetener of 1,2-benzoisothiazol-3-(2H)-one and is about 300-400 times sweeter than sucrose (55,56). It is less soluble in water in its natural form and hence it is conjugated with either sodium or calcium salt to improve on the solubility (55,56). Even though saccharin has not been reported to be tasted differently, research on various artificial sweeteners has reported that they may elicit different responses upon interaction with the sweet taste receptor, and as such, there is therefore a possibility of people having different tasting thresholds for saccharin (57).

The sensitivity testing protocol was adopted from 3M Center for Respiratory Protection, United States (58). While preparing for the testing, efforts were made to ensure that the study volunteers did not eat or drink anything, 30 minutes before the testing except for drinking water. The testing hood was assembled by fitting it onto the collar of the study volunteers, making a 10 cm ‘gap’ between the study volunteers’ faces and the inside surface of the hood. The nebulizers were prepared and about 10 mls of saccharin sensitivity and test solutions were added to their respective nebulizers. Sensitivity testing and test experiments were then performed. The sensitivity testing was performed to ensure that the study volunteers could taste the saccharin solution before carrying out the test experiments. Before nebulizing, the study volunteers were instructed to breathe through the mouth with the tongue slightly out, so that they could be able to taste the saccharin. During the fit testing, the study volunteers were instructed to breathe: (i) normally, (ii) deeply, (iii) while moving head side-to-side, (iv) while moving head up and down, (v) while bending over or bending at the waist, (vi) while talking, and (vii) to breathe normally again. When saccharin was not tasted by the study volunteers; the test was considered as ‘passed’ or ‘successful’ while the test was considered as ‘failed’ or ‘unsuccessful’ when the saccharin was tasted by the study volunteers. Despite, the ‘failed’ test, the number of squeezes applied for the study volunteers to taste the saccharin were recorded. The test was recorded as completely ‘failed’ or ‘unsuccessful’ after repeat testing had been done and the saccharin was tasted while the same face-mask was being worn.

### In-house filtration efficiency

In the testing, a 0.5 McFarland bacterial suspension of *Mycobacterium smegmatis* was prepared and later transferred into a multi-purpose hand-held sprayer that was used to generate a steady stream of aerosols. A vacuum filtration unit was constructed in-house and set up in a biosafety cabinet. All items in the biosafety cabinet were thoroughly disinfected using 5% Lysol and 70% ethanol. The in-built filter of the vacuum filtration unit was aseptically removed and replaced with the test filters (sections of the face-masks). The negative control protocol was performed by running the vacuum pump for 30 minutes without aerosol generation and the filtered air was inoculated on 7H11 agar media. The different face-masks were tested by maintaining a steady stream of aerosols of *Mycobacterium smegmatis* for 30 minutes directed towards the vacuum filtration unit having the face-mask sections and the filtered air was inoculated on 7H11 agar media. The positive control protocol was performed by maintaining a steady stream of aerosols of *Mycobacterium smegmatis* for 30 minutes without any filtration and inoculating the air/aerosols on 7H11 agar media. The inoculated agar plates were sealed with micropore tape and incubated at 37°C for 3 days. The filtration efficiency was assessed by comparing the growth on the plates for the positive control with those of the test face-masks.

### In-house breathability testing

In the testing, the ASTM E96 water vapor testing protocol specifically the desiccant method was adapted (59). In the testing, a glass beaker was almost filled with a solid desiccant (anhydrous calcium chloride granules) and then covered with the sections of the test face-masks leaving a very small air space between the desiccant and the sections of the test face-masks. The sections of the test face-masks were properly sealed to the edge of the glass beaker to prevent side diffusion. The initial weight of the glass beaker containing the calcium chloride granules was then taken. The glass beaker was then placed in an environment where the environmental -humidity and -temperature were constantly monitored and recorded. The glass beaker containing the calcium chloride granules was then weighed after 2 hours and the gain in weight of the set-up was extrapolated to provide data on the moisture passing through the fabric in a day (i.e. 24 hours). A positive control glass beaker containing calcium chloride was included. This was left uncovered for the entire duration of the experiment. The permeability of the face-masks was categorized as ‘low’, ‘moderate’ or ‘high’.

### In-house microbial cleanliness testing

This testing was performed after the study volunteers had used each of the face-mask for the fitness testing. After the fitness testing, each of the face-masks was aseptically removed from the study volunteers faces and the outer and inner surfaces of each of the used face-masks were swabbed in that order with sterile cotton swabs made slightly wet with physiological saline while focusing on the sites where the nose and mouth would typically be positioned. The contents of the swabs were then inoculated onto Blood agar and MacConkey agar. Both culture media were then incubated aerobically, at 35°C ± 2°C for 24 hours. All growth was enumerated and identified per the face-mask tested and the results recorded. The identification of target gram-positive bacteria (i.e. *Staphylococcus spp*) was done using recommended biochemical tests: DNase, mannitol salt fermentation after performing gram-stain microscopy, catalase, slide and tube coagulase tests.

### In-house distance dependent fit testing

In this testing: (i) care was taken to ensure that the study volunteers had no facial hair and that they did not eat or drink anything except drinking water in the 30 minutes before the start of the testing, (ii) the nebulizers (hand-held sprayers) were prepared and saccharin sensitivity and test solutions were added to their respective nebulizers, (iii) sensitivity testing was performed with the study volunteers not wearing face-masks, while standing stationery at 2, 3, 4, 5 and 6 meters, to ensure that the study volunteers could taste the saccharin sensitivity solution at the different distances. At distances and beyond which the saccharin sensitivity solution was not tasted by the study volunteers, testing with the saccharin test solution was not performed. The saccharin test solution was tested after the study volunteers wore the test face-masks at stood stationery at the distances where the saccharin sensitivity solution had been tasted by the same individuals. During the testing, particularly before nebulizing, the study volunteers were instructed to breathe through the mouth with the tongue slightly out, so that they could ably taste the saccharin. During the testing, the volunteers were instructed to breathe: (i) normally, (ii) deeply, (iii) while moving their head side-to-side, (iv) while moving their head up and down, (v) while bending over or bending at the waist, (vi) while talking, (vii) and to breathe normally again. When saccharin was not tasted at the respective distances; the test was considered as ‘passed’ or ‘successful’ while the test was considered as ‘failed’ or ‘unsuccessful’ when the saccharin was tasted at the respective distances. Despite, the ‘failed’ test, the number of squeezes to tasting the saccharin were recorded. The test was recorded as completely ‘failed’ or ‘unsuccessful’ after repeat testing was done and the saccharin was tasted while the same face-mask were being worn.

### In-house face-mask re-usability testing

In the testing, each of the face-masks was subjected to 3 decontamination methods that included: (i) spraying the face-masks with 70% ethanol and drying under the sun particularly for the non-reusable face-masks, (ii) washing the face-masks with non-bacterial soap and drying them under the sun, (iii) and, washing the face-masks with non-bacterial soap, drying them under the sun and ironing them, after which semi-quantitative mask fitness testing, in-house: face-mask filtration efficiency testing, breathability testing, and distance dependent fit testing were repeated.

### Positive controls

United States Food and Drug Administration (FDA) KN95 paper based European Union (EU) certified EN 149:2001+A1:2009 FFP2 NR D face-masks were used as positive controls in the series of experiments performed.

### Data management and analysis

Data from the collection forms previously validated by the study principal investigators and the laboratory team was entered and cleaned using MS Excel 2016 and analyzed using Stata 14.0, a statistical software. Descriptive analyses such as proportions, and means (where appropriate) were performed. In-house face-mask filtration efficiency was determined by counting the number of colony forming units (CFUs) on the test culture plates similar to those on the positive control culture plates. Analysis of variance was used to assess the association between the face-mask types and average increase in weight and humidity to determine which face-mask had the best breathability. The statistical significance levels were two-sided at p<0.05 (95% CI). Graphs were plotted using excel and Stata 14.0 where appropriate to describe the data.

## Data availability

The datasets generated and/or analyzed during the current study are not publicly available but are available from the corresponding author on reasonable request.

## Declarations

### Acknowledgement

We thank Okello Isaac Opio, Sserubiri James, Kaddu Arafat, and Musanje Isaac who volunteered for the laboratory testing, and Nagawa Bridget Tamale from Elevate Research Services, who helped in data analysis.

## Authors’ contributions

GM conceived and designed the study, provided technical guidance, supervised the work, reviewed the final manuscript. DS and HN performed the laboratory testing and participated in interpretation of the data, writing and manuscript reviews. DA conceived and designed the study, provided technical guidance, supervised the work, participated in analysis and interpretation of the data and drafted the manuscript. DB participated in data analysis and interpretation, writing and manuscript reviews. All the authors reviewed and approved the final manuscript.

## Additional information

### Correspondence and requests for materials should be addressed to D.A

#### Ethics approval and consent to participate

Ethical approvals were obtained from the: (1) School of Biomedical Sciences-Research and Ethics Committee, College of Health Sciences, Makerere University (approval number: SBS-793) and the Uganda National Council for Science and Technology (approval number: SS489ES). Written informed consent was obtained from each of the study volunteers before performing the laboratory investigations.

#### Funding

This work was supported by the Government of Uganda through Makerere University Research and Innovations Fund (RIF) under the RIF Special COVID-19 Research and Innovation Awards-2020 (Grant #: MAK/DVCFA/151/20). The views expressed herein are those of the author(s) and not necessarily those of the Government of Uganda and Makerere University Research and Innovations Fund.

